# Association of Antihypertensive Medication and Steatotic Liver Disease with Liver Fibrosis and Mortality among US Adults

**DOI:** 10.1101/2025.05.20.25328037

**Authors:** Yu Wu, Fei Fang

## Abstract

**Background:** Metabolic dysfunction-associated steatotic liver disease (MASLD) and hypertension frequently coexist in adults across the Americas, yet evidence to guide pharmacologic management in individuals with both conditions remains limited. Antihypertensive medications may influence liver outcomes, but comparative data across drug classes are sparse.

**Methods:** We performed a cross-sectional analysis using pooled data from the US National Health and Nutrition Examination Survey (1999 to 2018), linked to mortality records through 2019. Hepatic steatosis was assessed using the US Fatty Liver Index (US-FLI), Fatty Liver Index (FLI), and Hepatic Steatosis Index (HSI). Fibrosis was assessed using noninvasive scores. Antihypertensive exposure, identified through prescription records, included ACE inhibitors, ARBs, beta blockers, calcium channel blockers (CCBs), and diuretics. Associations with liver fibrosis were estimated using logistic regression. All cause and cardiovascular mortality were assessed using Cox proportional hazards models with inverse probability of treatment weighting.

**Findings:** Among 2,909 adults with MASLD receiving monotherapy antihypertensive treatment, use of ACEIs and ARBs was associated with lower odds of liver fibrosis compared with CCBs. In adjusted models, ACEIs were associated with reduced all-cause mortality (adjusted hazard ratio 0.30; 95% CI 0.11-0.82), as were ARBs (0.25; 95% CI 0.07-0.94). No significant differences were observed for cardiovascular mortality across medication classes.

**Interpretation:** Use of angiotensin converting enzyme inhibitors and angiotensin receptor blockers was associated with lower fibrosis burden and improved survival in individuals with metabolic dysfunction associated steatotic liver disease. These findings are hypothesis generating and warrant confirmation in prospective studies.

**Funding:** No Funding.

## Introduction

Metabolic dysfunction-associated steatotic liver disease (MASLD) has become a major public health concern, affecting an estimated 25% to 33% of the global population^1^. As the burden of viral hepatitis declines, MASLD has emerged as the leading cause of cirrhosis and hepatocellular carcinoma, particularly in the Americas^2,3^. The rising prevalence of obesity, type 2 diabetes, and hypertension has further accelerated the spread of MASLD, and projections indicate a continued increase in disease burden through 2030^4^.

MASLD is closely linked to metabolic syndrome, and a substantial proportion of individuals progress to metabolic dysfunction-associated steatohepatitis (MASH)^5^. MASH-related cirrhosis is now one of the most common indications for liver transplantation and a recognized risk factor for hepatocellular carcinoma^6^. Among the components of metabolic syndrome, hypertension plays a key role in worsening MASLD^7^. It promotes hepatic insulin resistance and increases fatty acid flux to the liver, partly through the effects of angiotensin II^8^. Preclinical studies have suggested that angiotensin-converting enzyme inhibitors (ACEIs) and angiotensin receptor blockers (ARBs), which inhibit the renin-angiotensin system, may attenuate hepatic fibrosis and inflammation^9,10^.

Given the high prevalence of MASLD and hypertension in populations across the Americas, especially in regions with elevated rates of metabolic comorbidities, evaluating the association between antihypertensive medications and liver-related outcomes is of clinical and public health importance^11^. In this study, we used data from the National Health and Nutrition Examination Survey (NHANES) to examine whether the use of ACEIs or ARBs is associated with liver fibrosis and all-cause mortality among adults with MASLD. These findings may contribute to risk stratification and therapeutic decision-making in the management of MASLD within primary care and public health settings.

## Methods

### Study population

We conducted a mortality-linked, retrospective cohort study using data from the National Health and Nutrition Examination Survey (NHANES), a nationally representative survey of the noninstitutionalized US population. NHANES collects demographic, socioeconomic, dietary, and health-related information through structured interviews, along with clinical, laboratory, and anthropometric assessments conducted in mobile examination centers. This study included adults aged 20 years and older who participated in NHANES from 1999 through 2018. The survey follows a multistage, stratified probability sampling design, with assigned sampling weights accounting for nonresponse and survey complexity.

Mortality outcomes were determined through linkage with the National Death Index (NDI), with follow-up available through December 31, 2019. All NHANES protocols were approved by the National Center for Health Statistics Institutional Review Board, and written informed consent was obtained from all participants. This analysis adheres to the Strengthening the Reporting of Observational Studies in Epidemiology (STROBE) guidelines.

### Participant Selection

Participants were eligible for inclusion if they were aged ≥20 years and had evidence of hepatic steatosis at baseline, defined by at least one of the following validated non-invasive indices: US Fatty Liver Index (US-FLI) ≥30^12^, Fatty Liver Index (FLI) ≥60^13^, or Hepatic Steatosis Index (HSI) ≥36^14^. These indices have been widely used in large epidemiologic studies for screening hepatic steatosis^15^.

Eligible participants were also required to have reported the use of at least one antihypertensive medication, categorized into one of the following classes: angiotensin receptor blockers (ARBs), angiotensin-converting enzyme inhibitors (ACEIs), beta blockers, calcium channel blockers (CCBs), or diuretics.

### Exclusion Criteria

Participants were excluded if they had a known history of viral hepatitis, including hepatitis B or C, or if they had missing data for steatosis assessment. Individuals with unknown alcohol use in the past 12 months, or missing information on antihypertensive medication use or mortality status, were also excluded. These criteria were applied to reduce potential misclassification and to isolate cases of steatosis likely attributable to metabolic dysfunction. After applying all inclusion and exclusion criteria, the final analytical cohort consisted of 2,909 participants.

### MASLD Classification

Metabolic dysfunction-associated steatotic liver disease (MASLD) was defined according to the 2023 international consensus criteria established by the Liver Disease Nomenclature Global Consensus Group^16^. MASLD includes individuals with hepatic steatosis in the presence of at least one cardiometabolic risk factor, after exclusion of secondary causes of liver disease such as viral hepatitis and excessive alcohol intake. This classification aims to distinguish MASLD from alcohol-associated liver disease (ALD) and metabolic dysfunction and alcohol-associated liver disease (MetALD), and to focus on liver injury primarily driven by metabolic dysregulation^17^.

Cardiometabolic risk factors used for MASLD classification included: (1) BMI ≥23 kg/m² or waist circumference ≥90 cm for males or ≥80 cm for females; (2) fasting glucose ≥5.6 mmol/L (100 mg/dL), type 2 diabetes, or the use of glucose-lowering medications; (3) blood pressure ≥130/85 mmHg, or the use of blood pressure-lowering medications; (4) triglycerides ≥1.70 mmol/L (150 mg/dL), or the use of lipid-lowering medications; (5) HDL cholesterol <1.0 mmol/L (40 mg/dL) for males or <1.3 mmol/L (50 mg/dL) for females, or the use of lipid-lowering medications.

Other clinical definitions were based on standard criteria. Type 2 diabetes (T2DM) was defined by an A1c ≥6.5%, fasting glucose ≥126 mg/dL, a self-reported diagnosis, or the use of glucose-lowering agents or insulin^18^. Hypertension was defined as a clinical diagnosis or systolic blood pressure ≥140 mmHg or diastolic pressure ≥90 mmHg, or the use of antihypertensive medications^19^. Hyperlipidemia was defined as serum cholesterol ≥200 mg/dL, LDL ≥130 mg/dL, HDL <40 mg/dL for men or <50 mg/dL for women, or a history of hyperlipidemia^20^.

This approach ensured consistency with previous research and aligned with contemporary international definitions, allowing for valid assessment of associations between MASLD and antihypertensive medication use in a nationally representative cohort.

### Exposure assessment

The primary outcome was liver fibrosis, assessed using noninvasive methods. Fibrosis classification in the main analysis was based on a predictive modeling framework, consistent with recommended two-tier strategies for liver fibrosis risk stratification^21^. Vibration-controlled transient elastography (VCTE) data from the 2017-2018 NHANES cycle served as the reference standard for model development. In this VCTE cohort, fibrosis was defined as a liver stiffness measurement (LSM) of 7.5 kPa or higher and a controlled attenuation parameter (CAP) of at least 268 dB/m^22^.

To enable fibrosis classification across earlier NHANES cycles, logistic regression models were trained to predict VCTE-defined fibrosis using routinely available serum-based indices: Fibrosis-4 index (FIB-4), NAFLD Fibrosis Score (NFS), AST to Platelet Ratio Index (APRI), and the BARD score^23^. Model performance was evaluated using the area under the receiver operating characteristic curve (AUROC), and optimal probability thresholds were determined using Youden’s J statistic. Participants with predicted probabilities above the model-derived cutoffs were classified as having liver fibrosis.

These model-based predictions were applied retrospectively to NHANES participants from 1999 to 2016, allowing for fibrosis classification across the full analytic cohort. In sensitivity analyses, individual noninvasive indices, including FIB-4, NFS, and APRI, were also examined separately using conventional thresholds to evaluate the robustness of fibrosis-outcome associations.

To reduce the potential for confounding due to overlapping antihypertensive therapies, we restricted the primary analysis to individuals receiving monotherapy. This approach facilitated more direct attribution of treatment effects to specific drug classes and reduced the risk of interaction bias. It also enabled estimation of stabilized propensity scores using multinomial logistic regression, improving covariate balance and model transparency. Although this restriction reduced the sample size, it strengthened internal validity and improved comparability across exposure groups.

### Mortality status

Mortality data were obtained through linkage with the National Death Index (NDI), with follow-up available through December 31, 2019. All-cause mortality was defined as death from any cause. Cardiovascular disease (CVD) mortality was defined based on the underlying cause of death, using the International Statistical Classification of Diseases and Related Health Problems, Tenth Revision (ICD-10). CVD mortality included deaths attributed to cardiovascular conditions (ICD-10 codes I00-I09, I11, I13, I20-I51) and cerebrovascular disease (ICD-10 codes I60-I69).

### Statistical analysis

Statistical analyses were conducted in a pooled cross-sectional cohort drawn from NHANES 1999-2018. Hazard ratios (HRs) with 95% confidence intervals (CIs) for all-cause and cardiovascular mortality were estimated using Cox proportional hazards models. Antihypertensive medication class was the primary exposure, with calcium channel blockers (CCBs) serving as the reference group. Separate models were constructed for participants with steatotic liver disease and for those who met MASLD criteria.

To adjust for baseline confounding across multiple treatment classes, inverse probability of treatment weights (IPTWs) were derived from a multinomial logistic regression model predicting the probability of receiving each medication class^24^. Covariates included age, sex, race and ethnicity, marital status, smoking status, family income-to-poverty ratio, body mass index, waist circumference, diabetes, hypertension, and hyperlipidemia^25,26^. Stabilized IPTWs were combined with NHANES examination weights in the outcome models to account for the complex survey design and ensure national representativeness.

Associations between antihypertensive class and liver fibrosis were examined using logistic regression. Analyses were conducted using both observed fibrosis status and model-predicted outcomes. In the 2017-2018 NHANES cycle, fibrosis was defined using VCTE-based thresholds. Logistic regression models were trained to predict fibrosis status using noninvasive indices including FIB-4, NAFLD fibrosis score (NFS), AST to platelet ratio index (APRI), and BARD score. Model performance was assessed using area under the receiver operating characteristic curve (AUROC), and optimal classification thresholds were selected using Youden’s J statistic, which maximizes the sum of sensitivity and specificity.

CCBs were selected as the reference category based on the highest unadjusted mortality and their stable behavior across models. Sensitivity analyses included alternative reference groups, outcome models without IPTW adjustment, and stratified models by sex, diabetes status, and income-to-poverty ratio^27,28,29^. Additional robustness checks were conducted in two phases. The first phase reassessed the relative associations of ACEIs and ARBs using alternate reference categories. The second phase re-estimated IPTWs using an expanded multinomial model with additional covariates^30^. Both phases yielded results consistent with the primary analysis. Interaction effects were tested using multiplicative terms and Wald statistics. All analyses incorporated the combined IPTW and NHANES weights. Statistical significance was defined as a two-sided P value less than 0.05. Analyses were conducted using R software (version 2023.09.1).

## Results

### Study population baseline characteristics

The final study cohort included 19,603 US adults from NHANES 1999-2018 (Supplementary Figure 1). The mean age was 49.5 years (SD 18.1), and 52.5% were women. Participants were predominantly non-Hispanic White (40.3%), with 16.5% non-Hispanic Black and 5.4% non-Hispanic Asian. Over the follow-up period, 1,045 cardiovascular deaths were recorded. Antihypertensive medication use was distributed as follows: 2,328 individuals used ACE inhibitors, 1,039 used ARBs, 2,255 used β-blockers, 1,283 used CCBs, and 1,928 used diuretics (Table 1).

**Table 1.** Baseline characteristics of participants by antihypertensive medication use.

| characteristics | No antihypertensive medication intake<br>(N=13889) | Antihypertensive medication intake<br>(N= 5714) |
| --- | --- | --- |
| Age, mean (SD), y | 43 (17) | 64 (13) |
| Sex (%) |  |  |
| Male | 6682 (48.1) | 2620 (45.9) |
| Female | 7207 (51.9) | 3094 (54.1) |
| Waist circumference, cm | 96 (15) | 106 (16) |
| Body mass index | 28 (6) | 31 (7) |
| Race/ethnicity |  |  |
| Non-Hispanic White | 5472 (39.4) | 2430 (42.5) |
| Non-Hispanic Black | 1985 (14.3) | 1254 (21.9) |
| Non-Hispanic Asian | 822 (5.9) | 237 (4.1) |
| Hispanic | 4993 (35.9) | 1600 (28.0) |
| Other or multiracial | 617 (4.4) | 193 (3.4) |
| Education level |  |  |
| less than high school graduate | 3482 (25.1) | 1719 (30.1) |
| High school graduate or GED | 3020 (21.8) | 1404 (24.6) |
| Some college or above | 7373 (53.1) | 2586 (45.3) |
| Marital status |  |  |
| Married or Living with Partner | 8586 (62.5) | 3432 (60.4) |
| Divorced or Living without Partner | 5154 (37.5) | 2249 (39.6) |
| Family income-to-poverty ratio | 2.6 (1.6) | 2.6 (1.6) |
| Laboratory examination results |  |  |
| ALT, U/L | 25 (26) | 24 (15) |
| AST, U/L | 25 (19) | 25 (16) |
| γGT, U/L | 27 (39) | 33 (40) |
| Triglycerides, mg/dL | 126 (123) | 148 (120) |
| Glucose, mmol/L | 103 (30) | 121 (44) |
| Insulin, uU/mL | 12 (14) | 17 (21) |
| HbA1c, % | 6 (1) | 6 (1) |
| HDL, mg/dL | 54 (16) | 53 (16) |
| Hypertension |  |  |
| Yes | 786 (5.7) | 5714 (100.0) |
| No | 13103 (94.3) | 0 (0.0) |
| Diabetes mellitus |  |  |
| Yes | 1060 (7.3) | 1827 (30.6) |
| No | 12825 (92.7) | 3886 (69.4) |
| Hyperlipidemia |  |  |
| Yes | 7153(51.5) | 4325 (75.7) |
| No | 6735(48.5) | 1389 (24.3) |
| Smoking status |  |  |
| Yes | 6022 (43.4) | 2833 (49.6) |
| No | 7854 (56.6) | 2878 (50.4) |
| Alcohol use |  |  |
| Never/light drinkers | 4255 (46.4) | 1903 (62.7) |
| Heavy drinkers | 4910 (53.6) | 1132 (37.3) |

Abbreviations: ALT, alanine aminotransferase; AST, aspartate aminotransferase; γGT, $\gamma$ -glutamyltransferase; HbA1c, Glycated Hemoglobin; HDL, high density lipoprotein. y represents year.
Values are presented as mean (SD) or number (%), unless otherwise specified.

Compared to non-users, individuals receiving antihypertensive medications were older, had higher BMI and waist circumference, and had greater prevalence of diabetes and hyperlipidemia. These findings reflect a higher burden of cardiometabolic risk and underscore the importance of evaluating their potential hepatic and survival benefits in MASLD.

### Association between noninvasive fibrosis tools and antihypertensive treatment

Figure 1 and Table 2 present the associations between antihypertensive medication use and advanced fibrosis, as defined by FIB-4, among adults with SLD and MASLD. In the US-FLI–defined population, use of antihypertensive medications was associated with significantly lower odds of fibrosis in individuals with SLD (adjusted hazard ratio [aHR], 0.36; 95% CI, 0.15-0.85) and MASLD (aHR, 0.20; 95% CI, 0.06-0.68), after adjustment using IPTW and NHANES sampling weights. No significant associations were observed in the FLI-defined cohort.

**Figure 1.**
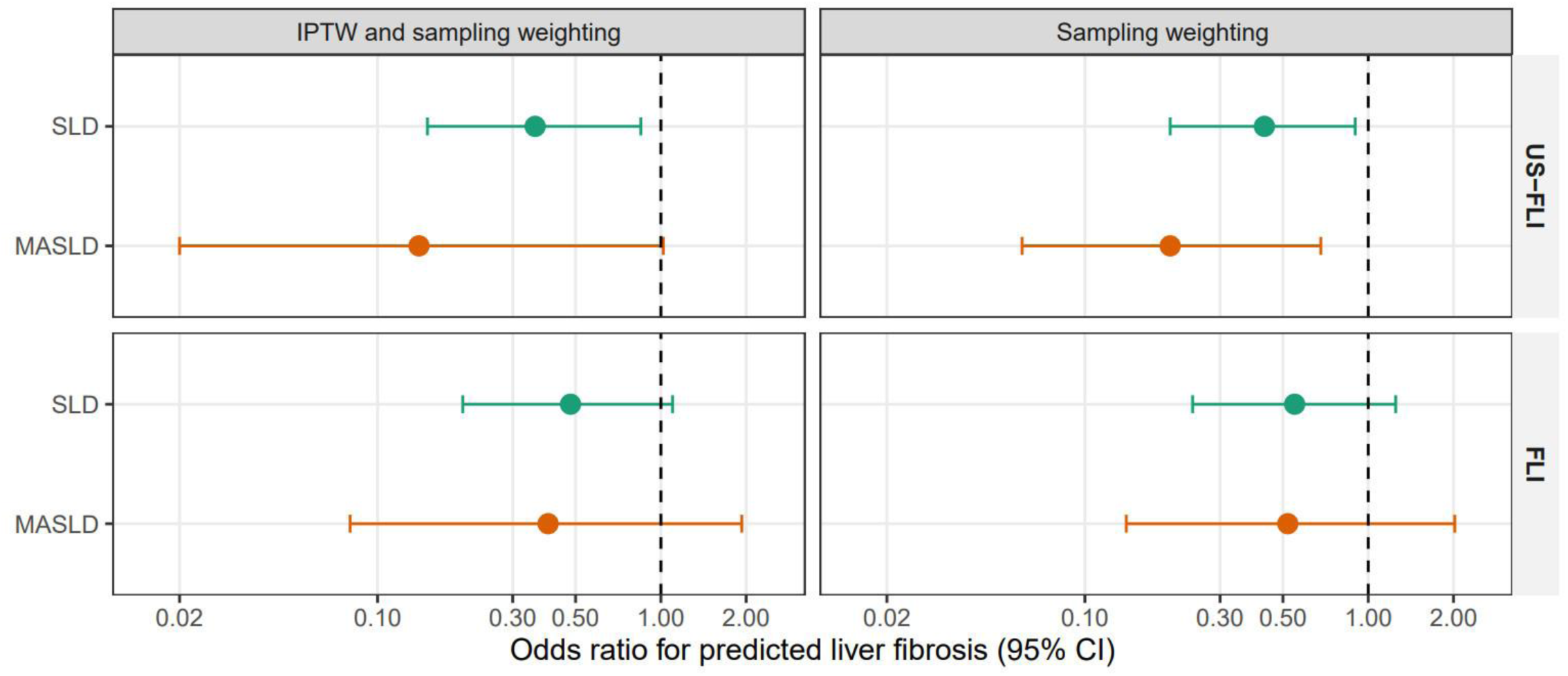
Associations between antihypertensive medication use and FIB-4–defined liver fibrosis. Adjusted odds ratios and 95% confidence intervals were estimated from logistic regression models evaluating the association between antihypertensive medication use and FIB-4–defined liver fibrosis. Models were stratified by steatosis classification (US-FLI and FLI) and adjusted using IPTW with sampling weights or sampling weights only. Full model results are presented in Table 2.

**Table 2.** Adjusted odds ratios for the association between antihypertensive medication use and FIB-4–defined liver fibrosis across different steatosis definitions.

|  |  |  | IPTW and Sampling Weighting |  |  | Only Sampling Weighting |  |  |
| --- | --- | --- | --- | --- | --- | --- | --- | --- |
| Population Defined by US-FLI | LF (FIB-4) NO. (%) | non-LF (FIB-4) No. (%) | Logistic regression coefficient | aOR (95% CI) | P-value | Logistic regression coefficient | aOR (95% CI) | P-value |
| SLD | 60 (1.43) | 4133 (98.57) | -1.03 | 0.36 (0.15-0.85) | 0.02 | -0.85 | 0.43 (0.20-0.90) | 0.03 |
| MASLD | 15 (1.16) | 1283 (98.84) | -1.96 | 0.14 (0.02-1.02) | 0.052 | -1.60 | 0.20 (0.06-0.68) | 0.01 |
| Population Defined by FLI | LF (FIB-4) NO. (%) | non-LF (FIB-4) No. (%) | Coefficient of AHM | aOR (95% CI) | P-value | Coefficient of AHM | aOR (95% CI) | P-value |
| SLD | 56 (1.12) | 4955 (98.88) | -0.74 | 0.48 (0.20-1.10) | 0.08 | -0.59 | 0.55 (0.24-1.25) | 0.15 |
| MASLD | 11 (0.70) | 1556 (99.30) | -0.92 | 0.40 (0.08-1.93) | 0.25 | -0.64 | 0.52 (0.14-2.02) | 0.35 |

Abbreviations: FLI, fatty liver index; FIB-4, Fibrosis-4 index; LF, liver fibrosis; SLD, steatotic liver disease; MASLD, metabolic dysfunction-associated steatotic liver disease; aOR, adjusted odds ratio; AHM, antihypertensive medication; IPTW, inverse probability of treatment weighting; CI, confidence interval.
Adjusted odds ratios (aORs) and 95% confidence intervals (CIs) estimated using logistic regression models. Analyses were stratified by steatosis definition (US-FLI, FLI) and liver disease category (SLD, MASLD), and adjusted using IPTW with NHANES sampling weights or sampling weights only.

Consistent findings were observed when fibrosis was assessed using APRI in the US-FLI–defined population (aHR, 0.62; 95% CI, 0.41-0.94) and FIB-4 in the HSI-defined population (aHR, 0.32; 95% CI, 0.11-0.91) (Supplementary Tables 1 and 5). No significant associations were identified using NFS or BARD scores across definitions (Supplementary Tables 1-6).

The discriminatory performance of fibrosis markers was evaluated using AUROC. NFS and BARD demonstrated higher predictive accuracy (AUROC 0.74; 95% CI, 0.72-0.76), while FIB-4 and APRI showed lower but acceptable performance (AUROC 0.61; 95% CI, 0.58-0.63) (Supplementary Table 7).

### Association between mortality and antihypertensive medication

Over a mean follow-up of 9.75 years (SD 5.54), 3,260 deaths were recorded, including 1,045 due to cardiovascular disease. Adjusted Cox regression models are summarized in Tables 3 and 4. Among individuals with SLD, use of ACE inhibitors was associated with significantly lower all-cause mortality compared to CCBs in sampling weight-adjusted models (aHR 0.51; 95% CI, 0.30-0.81; Figure 2, Table 3).

**Figure 2.**
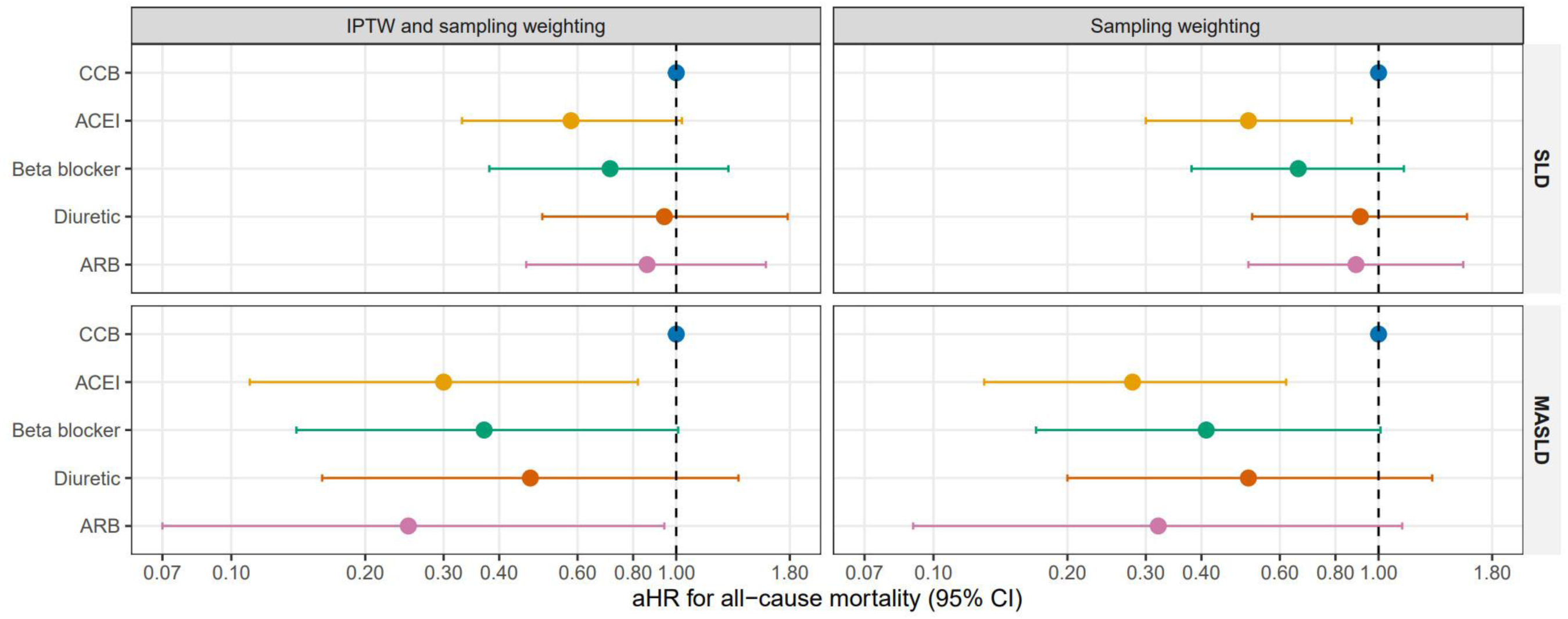
Associations between antihypertensive medication class and all-cause mortality among adults with SLD and MASLD. Adjusted hazard ratios and 95% confidence intervals were estimated using Cox proportional hazards models comparing each class of antihypertensive medication to CCBs. Separate models were constructed for individuals with SLD and MASLD, adjusted using IPTW and/or sampling weights. Full results are shown in Tables 3 and 4.

**Table 3.** Adjusted hazard ratios for all-cause mortality by antihypertensive medication class among participants with steatotic liver disease (SLD)

| (SLD, All-cause mortality) |  |  |  |  |  |  |  |
| --- | --- | --- | --- | --- | --- | --- | --- |
| AHM-class | IPTW and sampling weighting |  |  |  |  | Only sampling weighting |  |
|  | Patients NO.(%) | Event NO.(%) | PYs | aHR (95% CI) | P-value | aHR (95% CI) | P-value |
| CCB | 642 (13.65) | 208 (13.93) | 4801.33 | *Reference* |  | *Reference* |  |
| ACEI | 1273 (27.07) | 369 (24.72) | 10593.42 | 0.58 (0.33,1.03) | 0.06 | 0.51 (0.30,0.87) | 0.01 |
| Beta blocker | 1192 (25.35) | 395 (26.46) | 9079.92 | 0.71 (0.38,1.31) | 0.27 | 0.66 (0.38,1.14) | 0.14 |
| Diuretic | 1043 (22.18) | 377 (25.25) | 8397.42 | 0.94 (0.50,1.78) | 0.86 | 0.91 (0.52,1.58) | 0.73 |
| ARB | 552 (11.74) | 144 (9.65) | 4077.33 | 0.86 (0.46,1.59) | 0.62 | 0.89 (0.51,1.55) | 0.67 |

Abbreviations: SLD, steatotic liver disease; ACEI, angiotensin-converting enzyme inhibitor; ARB, angiotensin receptor blocker; CCB, calcium channel blocker; aHR, adjusted hazard ratio; AHM, antihypertensive medication; PYs, person-years; IPTW, inverse probability of treatment weighting; CI, confidence interval.
Estimates from Cox proportional hazards models comparing each antihypertensive class to calcium channel blockers (CCBs) as reference.
Models adjusted for age, sex, race/ethnicity, marital status, BMI, waist circumference, diabetes, hypertension, hyperlipidemia, smoking status, and income-to-poverty ratio, using IPTW with sampling weights or sampling weights only.

**Table 4.**
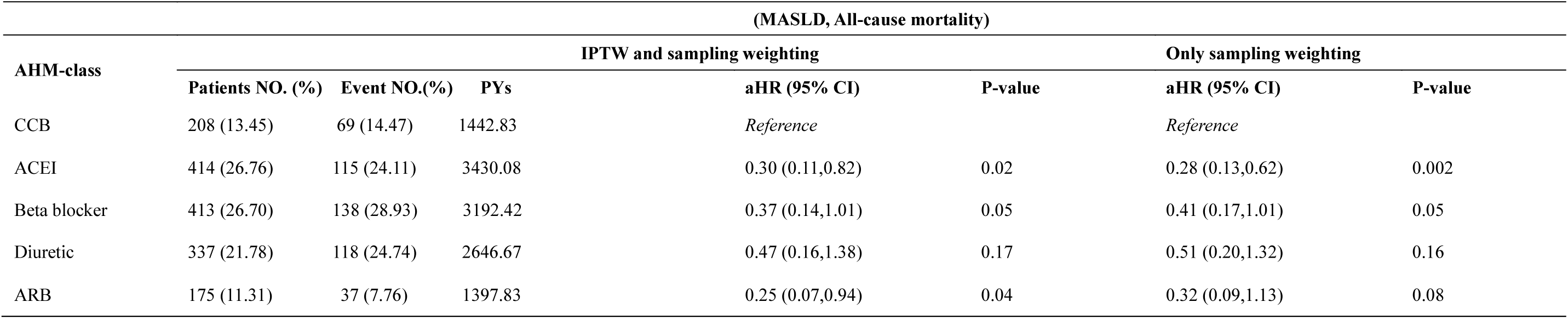
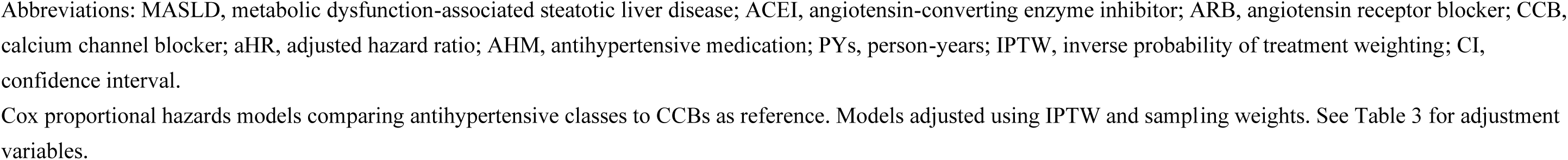
Adjusted hazard ratios for all-cause mortality by antihypertensive medication class among participants with MASLD.

| (MASLD, All-cause mortality) |  |  |  |  |  |  |  |
| --- | --- | --- | --- | --- | --- | --- | --- |
| AHM-class | IPTW and sampling weighting |  |  |  |  | Only sampling weighting |  |
|  | Patients NO. (%) | Event NO.(%) | PYs | aHR (95% CI) | P-value | aHR (95% CI) | P-value |
| CCB | 208 (13.45) | 69 (14.47) | 1442.83 | *Reference* |  | *Reference* |  |
| ACEI | 414 (26.76) | 115 (24.11) | 3430.08 | 0.30 (0.11,0.82) | 0.02 | 0.28 (0.13,0.62) | 0.002 |
| Beta blocker | 413 (26.70) | 138 (28.93) | 3192.42 | 0.37 (0.14,1.01) | 0.05 | 0.41 (0.17,1.01) | 0.05 |
| Diuretic | 337 (21.78) | 118 (24.74) | 2646.67 | 0.47 (0.16,1.38) | 0.17 | 0.51 (0.20,1.32) | 0.16 |
| ARB | 175 (11.31) | 37 (7.76) | 1397.83 | 0.25 (0.07,0.94) | 0.04 | 0.32 (0.09,1.13) | 0.08 |

Abbreviations: MASLD, metabolic dysfunction-associated steatotic liver disease; ACEI, angiotensin-converting enzyme inhibitor; ARB, angiotensin receptor blocker; CCB, calcium channel blocker; aHR, adjusted hazard ratio; AHM, antihypertensive medication; PYs, person-years; IPTW, inverse probability of treatment weighting; CI, confidence interval.

In the MASLD subgroup, ACE inhibitors were consistently associated with lower mortality in both IPTW-weighted models (aHR 0.30; 95% CI, 0.11-0.82) and sampling weight–adjusted models (aHR 0.28; 95% CI, 0.13-0.62). ARBs were also associated with a lower risk of all-cause mortality in IPTW-weighted analyses (aHR 0.25; 95% CI, 0.07-0.94; Figure 2, Table 4).

No significant associations were observed between antihypertensive medication class and cardiovascular-specific mortality in either SLD or MASLD populations (Supplementary Tables 9-10). Similarly, analyses in the FLI-defined population showed no significant associations between use of antihypertensive medications and all-cause mortality (Supplementary Table 8).

### Sensitivity and subgroup analyses

Sensitivity analyses using an alternative propensity score model yielded consistent findings. Among participants with MASLD, ACE inhibitors remained associated with significantly lower all-cause mortality compared to CCBs (aHR 0.31; 95% CI, 0.12-0.85), as did ARBs (aHR 0.25; 95% CI, 0.06-0.98) (Supplementary Table 15), supporting the robustness of the primary results (Table 4).

Subgroup analyses stratified by sex, income-to-poverty ratio, and baseline diabetes status revealed no significant interaction effects. Associations between ACEIs and all-cause mortality remained consistent across strata (Supplementary Tables 11-13).

Additional analyses using alternative reference groups showed that CCBs and diuretics were generally associated with higher mortality compared to ACE inhibitors. ACE inhibitors also showed a trend toward lower mortality compared to ARBs and diuretics (Supplementary Table 14), further supporting their potential benefit in MASLD.

## Discussion

In this nationally representative study of US adults, use of ACEIs and ARBs was associated with lower prevalence of liver fibrosis and reduced risk of all-cause mortality among individuals with steatotic liver disease (SLD) and metabolic dysfunction– associated steatotic liver disease (MASLD)^31,32^. These associations were consistent across multiple fibrosis assessment tools and confirmed in sensitivity analyses. The findings highlight a potential therapeutic role for renin-angiotensin system (RAS) inhibitors in improving long-term outcomes in patients with MASLD, a population with high cardiometabolic risk^33^. Interestingly, while ACEIs and ARBs were associated with lower all-cause mortality, no significant associations were observed for cardiovascular mortality. This discrepancy may reflect limited power to detect cardiovascular-specific effects due to fewer events. Alternatively, it may indicate that non-cardiovascular causes such as liver-related or oncologic mortality contributed more significantly to the observed survival benefit. Further studies with adjudicated cause-specific outcomes are needed to clarify this relationship.

Among the non-invasive tools applied to assess liver fibrosis, FIB-4 and APRI demonstrated sufficient predictive performance to detect clinically relevant associations despite their lower AUROC compared to NFS and BARD^33,34^. Their simplicity and availability in routine clinical data make them valuable for large-scale epidemiologic studies such as NHANES, where imaging or biopsy data are unavailable^35^. Our findings support the use of these indices for risk stratification in MASLD populations and provide additional validation of their utility in public health surveillance.

This study builds on emerging evidence that RAS blockade may attenuate fibrosis progression and reduce adverse outcomes in patients with fatty liver disease. Prior analyses in advanced NAFLD or cirrhosis cohorts have reported benefits of ACEIs and ARBs in reducing portal pressure, liver-related decompensation, and mortality^36,37,38^. A recent study by Ng and colleagues using TriNetX demonstrated survival benefit and reductions in liver-related events among MASLD patients receiving RAS inhibitors^9^. However, that study primarily included patients with established liver disease, with outcomes defined by administrative codes and hospitalizations. In contrast, our study focused on earlier-stage disease and used non-invasive fibrosis markers to evaluate risk before clinical progression. Furthermore, we applied monotherapy-based exposure definitions and inverse probability of treatment weighting to reduce confounding from medication overlap, allowing for more precise comparisons between drug classes.

The survival benefit observed in our MASLD population may reflect both hepatic and extrahepatic effects of RAS inhibition. Mechanistically, ACEIs and ARBs have been shown to reduce hepatic stellate cell activation, oxidative stress, and inflammation^39^. These agents also improve insulin sensitivity and endothelial function, which may contribute to reductions in systemic mortality risk. Although we did not observe a significant association with cardiovascular-specific mortality, the consistent reduction in all-cause mortality suggests a broader metabolic benefit^40^. Notably, ACE inhibitors showed stronger and more consistent associations with mortality reduction than other classes.

From a regional health perspective, the co-occurrence of MASLD and hypertension is highly prevalent across the Americas^41^. Data suggest that more than half of MASLD patients in the United States have elevated blood pressure^42^. As ACEIs and ARBs are widely available and form the cornerstone of hypertension management, their potential benefit in MASLD offers an opportunity to align cardiovascular and liver disease strategies in primary care settings. These findings support integrated approaches in resource-limited environments, where non-invasive fibrosis screening and RAS inhibition may offer cost-effective options for managing multimorbidity^43,44^.

Our findings also carry implications for clinical practice. Primary care providers routinely manage patients with hypertension, diabetes, and obesity, which are key components of MASLD. Fibrosis scores such as FIB-4 and APRI can be calculated using standard laboratory values and may help identify high-risk patients who could benefit from therapies that offer both metabolic and hepatic benefits. Integrating fibrosis risk assessment into routine hypertension management may support early identification of individuals at risk, especially in underserved populations with limited access to specialty care.

Several limitations should be acknowledged when interpreting these findings. First, although NHANES provides nationally representative, mortality-linked data, its cross-sectional design limits the capacity to draw causal conclusions. Specifically, the use of cross-sectional data limits adjustment to baseline covariates and does not account for changes in treatment exposure, liver disease progression, or time-varying confounding. Consequently, models that adjust only for baseline factors have limited ability to support causal interpretation. Second, fibrosis was classified using non-invasive indices rather than histology or elastography, which may reduce diagnostic accuracy^45^. Third, alcohol use was based on self-report and may have introduced misclassification between MASLD, MetALD, and alcohol-associated liver disease^46^. Fourth, we restricted the primary analysis to individuals receiving antihypertensive monotherapy in order to isolate the effects of specific drug classes. This approach allowed for more precise exposure classification, reduced confounding from drug combinations, and enabled the use of multinomial logistic regression for propensity score estimation. Importantly, it improved model interpretability and precision in estimating the effects of individual treatments. However, this decision also limited the sample size and excluded potentially relevant patient groups who receive multiple treatments, and it may overlook interaction effects among medications, increasing the risk of model misspecification. In addition, modeling propensity scores for multiple treatments would require treating each combination of medications as a distinct exposure, which could induce high variance in the estimated coefficients for each exposure.

Finally, although censoring weights were not incorporated in the present analysis, their application in future longitudinal studies could improve the precision of treatment effect estimates by accounting for dropout and unequal follow-up duration.

To sum up, given the observational design of NHANES and the absence of longitudinal treatment exposure data, our findings have limited power to support causal conclusions. Although IPTW was employed to mitigate confounding and enhance comparability across treatment groups, residual bias from unmeasured or time-varying covariates may persist. Therefore, the observed associations between antihypertensive medication class and liver-related or mortality outcomes should be interpreted as hypothesis-generating rather than conclusive evidence of a treatment effect. Prospective cohort studies or pragmatic trials will be essential to validate these findings and better understand the temporal dynamics of medication exposure and disease progression.

These findings underscore the need for integrated management strategies that consider both cardiovascular and hepatic risk in patients with MASLD. Prospective studies incorporating medication adherence, longitudinal liver assessments, and adjudicated outcomes are essential next steps.

## Conclusion

In this nationally representative cohort of US adults, use of antihypertensive medications was associated with a lower risk of liver fibrosis among individuals with steatotic liver disease and metabolic dysfunction-associated steatotic liver disease. Among treatment classes, ACE inhibitors and angiotensin receptor blockers were consistently associated with reduced all-cause mortality in MASLD, suggesting a potential benefit beyond blood pressure control.

While these findings are hypothesis-generating, they should be interpreted with caution due to the cross-sectional nature of the data. The study does not establish whether antihypertensive use preceded the development of fibrosis, nor does it capture treatment duration, adherence, or dynamic changes in liver status.

While ACE inhibitors and ARBs are not currently indicated specifically for the management of MASLD, their favorable associations with both fibrosis markers and survival in this study suggest that they may offer dual benefits for cardiometabolic and liver outcomes. These findings support further evaluation in prospective cohort studies and pragmatic trials, particularly in primary care and resource-limited settings across the Americas, where the burden of MASLD and hypertension frequently converge.

## Abbreviations

ACEI: angiotensin-converting enzyme inhibitor
aHR: adjusted hazard ratio
ALD: alcohol-associated liver disease
APRI: aspartate aminotransferase to platelet ratio index
ARB: angiotensin receptor blocker
AST: aspartate aminotransferase
CCB: calcium channel blocker
CVD: cardiovascular disease
FIB-4: Fibrosis-4 index
FLI: fatty liver index
γGT: gamma-glutamyltransferase
HbA1c: glycated hemoglobin
HDL: high-density lipoprotein
HSI: hepatic steatosis index
IPTW: inverse probability of treatment weighting
LF: liver fibrosis
MASLD: metabolic dysfunction-associated steatotic liver disease
MetALD: MASLD with increased alcohol intake
NFS: NAFLD fibrosis score
NHANES: National Health and Nutrition Examination Survey
SLD: steatotic liver disease
US-FLI: US Fatty Liver Index

## Contributors

YW and FF conceptualized the study. FF conducted the statistical analyses. YW drafted the manuscript. Both authors contributed to result interpretation and manuscript revision. All authors had full access to the data and approved the final version of the manuscript for submission.

## Data sharing statement

The datasets used in this study are publicly available through the National Health and Nutrition Examination Survey (NHANES) portal at https://www.cdc.gov/nchs/nhanes/index.htm.

## Artificial intelligence (AI) use

ChatGPT-5 was used for grammar review.

## Data Availability

All data produced in the present study are available upon reasonable request to the authors

https://www.cdc.gov/nchs/nhanes/

## Acknowledgements

We sincerely acknowledge the contributions of NHANES participants, staff, and data collection teams.

## Ethics statement

All NHANES protocols were approved by the National Center for Health Statistics (NCHS) Research Ethics Review Board. Written informed consent was obtained from all participants.

## Declaration of interests

The authors declare no competing interests.

## Notes

### Competing Interest Statement

The authors have declared no competing interest.

### Funding Statement

This study did not receive any funding

### Summary of Updates

This version of the manuscript includes substantial textual and structural revisions based on further refinement of the analytic models and additional context for interpretation. The key updates are as follows: Abstract was revised for clarity and conciseness, with improved articulation of background, methods, and findings. Introduction section was streamlined to better frame the relevance of antihypertensive therapy in MASLD and its implications for regional public health. Main results were reorganized to present fibrosis and mortality associations by exposure group and liver disease classification, emphasizing consistency across definitions (US-FLI, FLI, HSI). Discussion was significantly revised to reflect emerging clinical implications, highlight strengths and limitations in light of cross-sectional data, and underscore the potential therapeutic role of RAS inhibitors in MASLD. Multiple figures and supplementary tables were updated to match the refined statistical output. These revisions were made to strengthen methodological rigor, clarify causal interpretations, and improve readability for the medRxiv audience.

