## Supplementary Material for "Association of Antihypertensive Medication and Steatotic Liver Disease with Liver Fibrosis and Mortality among US Adults"

Supplemental Figure 1. Flow diagram of participant selection and exclusion

Supplemental Table 1. Adjusted logistic regression models for the association between antihypertensive medication use and liver fibrosis in individuals with SLD defined by US-FLI

Supplemental Table 2. Adjusted logistic regression models for the association between antihypertensive medication use and liver fibrosis in individuals with MASLD defined by US-FLI

Supplemental Table 3. Adjusted logistic regression models for the association between antihypertensive medication use and liver fibrosis in individuals with SLD defined by FLI

Supplemental Table 4. Adjusted logistic regression models for the association between antihypertensive medication use and liver fibrosis in individuals with MASLD defined by FLI

Supplemental Table 5. Adjusted logistic regression models for the association between antihypertensive medication use and liver fibrosis in individuals with SLD defined by HSI

Supplemental Table 6. Adjusted logistic regression models for the association between antihypertensive medication use and liver fibrosis in individuals with MASLD defined by HSI

Supplemental Table 7. Predictive performance of logistic models using FIB-4, NFS, APRI, and BARD for VCTE-defined fibrosis

Supplemental Table 8. Adjusted Cox models for all-cause mortality in SLD and MASLD defined by US-FLI and FLI

Supplemental Table 9. Adjusted Cox models for cardiovascular mortality in individuals with SLD

Supplemental Table 10. Adjusted Cox models for cardiovascular mortality in individuals with MASLD

Supplemental Table 11. Subgroup analysis by sex: associations between AHM class and all-cause mortality in MASLD

Supplemental Table 12. Subgroup analysis by income: associations between AHM class and all-cause mortality in SLD

Supplemental Table 13. Subgroup analysis by diabetes status: associations between AHM class and all-cause mortality in MASLD

Supplemental Table 14. Sensitivity analysis in SLD: using alternative reference groups to compare AHM classes and all-cause mortality

Supplemental Table 15. Sensitivity analysis in MASLD: using alternative IPTW models to evaluate associations between AHM class and all-cause mortality

**Supplemental Figure 1. Flow diagram of participant selection and exclusion**

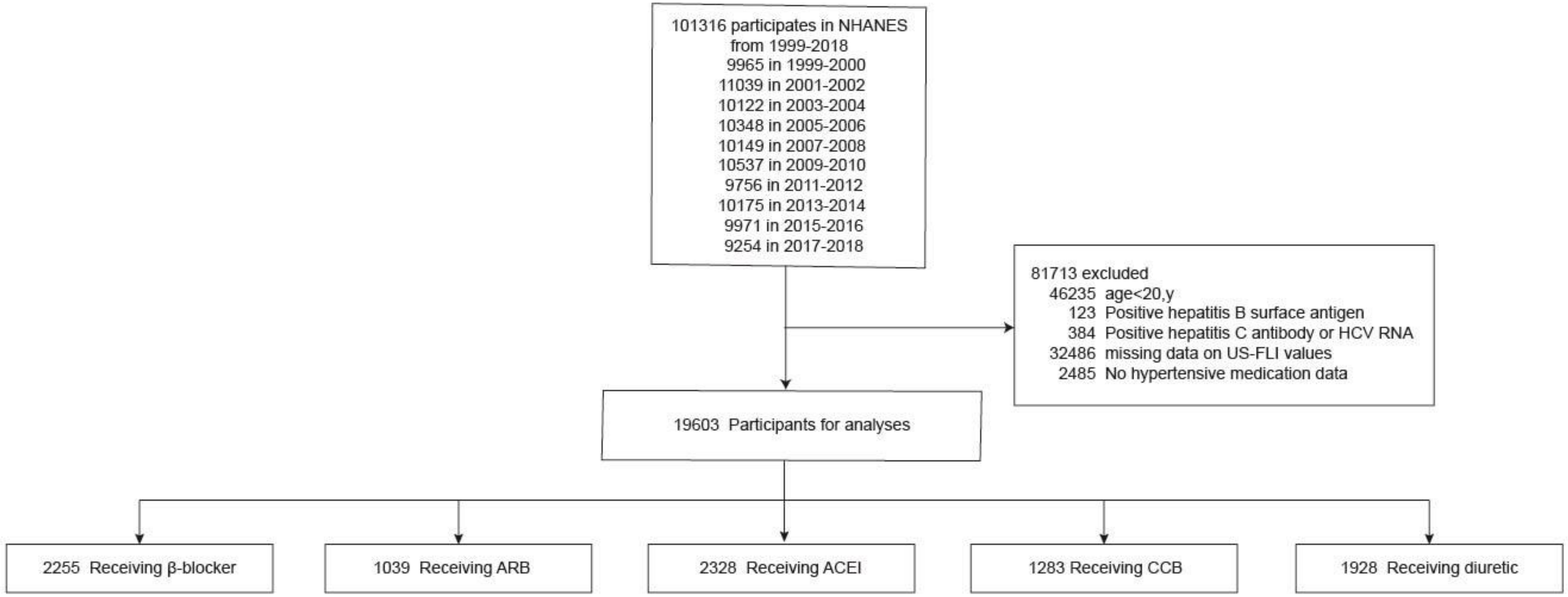


Abbreviations: ACEI, angiotensin-converting enzyme inhibitor; ARB, angiotensin receptor blocker; CCB, calcium channel blocker; NHANES, National Health and Nutrition Examination Survey; US-FLI, United States Fatty Liver Index.

Flow diagram depicting NHANES participant selection, exclusion criteria, and final analytical cohort for fibrosis and mortality analyses.

**Supplemental Table 1. Adjusted logistic regression models for the association between antihypertensive medication (AHM) use and liver fibrosis (LF) in individuals with SLD defined by US-FLI**

| | | | IPTW and Sampling weighting | | | Only Sampling Weighting | | |
|---|---|---|---|---|---|---|---|---|
| Outcomes | Cases (LF) NO. (%) | Controls (non-LF) No. (%) | Logistic regression coefficient | aOR (95% CI) | P-value | Logistic regression coefficient | aOR (95% CI) | P-value |
| FIB-4 | 60 (1.43) | 4133 (98.57) | -1.03 | 0.36 (0.15-0.85) | 0.02 | -0.85 | 0.43 (0.20-0.90) | 0.03 |
| NFS | 1268 (30.48) | 2892 (69.52) | -0.02 | 0.98 (0.68-1.42) | 0.92 | -0.08 | 0.92 (0.66-1.29) | 0.63 |
| APRI | 277 (6.61) | 3916 (93.39) | -0.48 | 0.62 (0.41-0.94) | 0.02 | -0.45 | 0.64 (0.43-0.94) | 0.02 |
| BARD | 1111 (26.37) | 3102 (73.63) | 0.22 | 1.25 (0.83-1.87) | 0.28 | 0.28 | 1.32 (0.90-1.95) | 0.16 |

Abbreviations: FIB-4, Fibrosis-4; NFS, NAFLD fibrosis score; APRI, aspartate aminotransferase to platelets ratio index; LF, liver fibrosis; SLD, steatotic liver disease; aOR, adjusted odds ratio; AHM, antihypertensive medication; IPTW, inverse probability of treatment weighting.

**Supplemental Table 2. Adjusted logistic regression models for the association between antihypertensive medication (AHM) use and liver fibrosis (LF) in individuals with MASLD defined by US-FLI**

| | | | IPTW and Sampling weighting | | | Only Sampling Weighting | | |
|---|---|---|---|---|---|---|---|---|
| Outcomes | Cases (LF) NO. (%) | Controls (non-LF) No. (%) | Logistic regression coefficient | aOR (95% CI) | P-value | Logistic regression coefficient | aOR (95% CI) | P-value |
| FIB-4 | 15 (1.16) | 1283 (98.84) | -1.96 | 0.14 (0.02-1.02) | 0.052 | -1.60 | 0.20 (0.06-0.68) | 0.01 |
| NFS | 395 (30.60) | 896 (69.40) | 0.36 | 1.43 (0.75-2.74) | 0.27 | -0.27 | 1.31 (0.74-2.32) | 0.35 |
| APRI | 72 (5.55) | 1226 (94.45) | -0.18 | 0.84 (0.39-1.80) | 0.65 | -0.25 | 0.78 (0.36-1.72) | 0.53 |
| BARD | 346 (26.41) | 964 (73.59) | -0.21 | 0.81 (0.42-1.55) | 0.53 | 0.00 | 1.00 (0.54-1.84) | 0.99 |

Abbreviations: FIB-4, Fibrosis-4; NFS, NAFLD fibrosis score; APRI, aspartate aminotransferase to platelets ratio index; LF, liver fibrosis; MASLD, metabolic dysfunction-associated steatotic liver disease; aOR, adjusted odds ratio; AHM, antihypertensive medication; IPTW, inverse probability of treatment weighting.

**Supplemental Table 3. Adjusted logistic regression models for the association between antihypertensive medication (AHM) use and liver fibrosis (LF) in individuals with SLD defined by FLI**

| | | | IPTW and Sampling weighting | | | Only Sampling Weighting | | |
|---|---|---|---|---|---|---|---|---|
| Outcomes | Cases (AF) NO. (%) | Controls (non-AF) No. (%) | Logistic regression coefficient | aOR (95% CI) | P-value | Logistic regression coefficient | aOR (95% CI) | P-value |
| FIB-4 | 56 (1.12) | 4955 (98.88) | -0.74 | 0.48 (0.20-1.10) | 0.08 | -0.59 | 0.55 (0.24-1.25) | 0.15 |
| NFS | 1423 (28.40) | 3587 (71.60) | 0.07 | 1.08 (0.77-1.51) | 0.68 | 0.01 | 1.01 (0.73-1.39) | 0.95 |
| APRI | 281 (5.61) | 4730 (94.39) | -0.37 | 0.69 (0.46-1.06) | 0.09 | -0.32 | 0.73 (0.49-1.07) | 0.11 |
| BARD | 1327 (26.37) | 3705 (73.63) | 0.19 | 1.21 (0.80-1.82) | 0.37 | 0.26 | 1.29 (0.87-1.92) | 0.20 |

Abbreviations: FIB-4, Fibrosis-4; NFS, NAFLD fibrosis score; APRI, aspartate aminotransferase to platelets ratio index; LF, liver fibrosis; SLD, steatotic liver disease; aOR, adjusted odds ratio; AHM, antihypertensive medication; IPTW, inverse probability of treatment weighting.

**Supplemental Table 4. Adjusted logistic regression models for the association between antihypertensive medication (AHM) use and liver fibrosis (LF) in individuals with MASLD defined by FLI**

| | | | IPTW and Sampling weighting | | | Only Sampling Weighting | | |
|---|---|---|---|---|---|---|---|---|
| Outcomes | Cases (LF) NO. (%) | Controls (non-LF) No. (%) | Logistic regression coefficient | aOR (95% CI) | P-value | Logistic regression coefficient | aOR (95% CI) | P-value |
| FIB-4 | 11 (0.70) | 1556 (99.30) | -0.92 | 0.40 (0.08-1.93) | 0.25 | -0.64 | 0.52 (0.14-2.02) | 0.35 |
| NFS | 431 (27.50) | 1136 (72.50) | 0.34 | 1.41 (0.77-2.57) | 0.27 | 0.29 | 1.34 (0.77-2.33) | 0.30 |
| APRI | 70 (4.47) | 1497 (95.53) | -0.13 | 0.88 (0.37-2.11) | 0.77 | -0.17 | 0.85 (0.35-2.03) | 0.71 |
| BARD | 401 (25.41) | 1177 (74.59) | -0.31 | 0.73 (0.38-1.42) | 0.36 | -0.14 | 0.87 (0.45-1.69) | 0.68 |

Abbreviations: FIB-4, Fibrosis-4; NFS, NAFLD fibrosis score; APRI, aspartate aminotransferase to platelets ratio index; MASLD, metabolic dysfunction-associated steatotic liver disease; LF, liver fibrosis; aOR, adjusted odds ratio; AHM, antihypertensive medication; IPTW, inverse probability of treatment weighting.

**Supplemental Table 5. Adjusted logistic regression models for the association between antihypertensive medication (AHM) use and liver fibrosis (LF) in individuals with SLD defined by HSI**

| | | | IPTW and Sampling weighting | | | Only Sampling Weighting | | |
|---|---|---|---|---|---|---|---|---|
| Outcomes | Cases (LF) NO. (%) | Controls (non-LF) No. (%) | Logistic regression coefficient | aOR (95% CI) | P-value | Logistic regression coefficient | aOR (95% CI) | P-value |
| FIB-4 | 42 (0.73) | 5704 (99.27) | -1.15 | 0.32 (0.11-0.91) | 0.03 | -1.02 | 0.36 (0.12-1.05) | 0.06 |
| NFS | 1513 (26.33) | 4233 (73.67) | 0.06 | 0.06 (0.76-1.48) | 0.72 | -0.01 | 0.99 (0.72-1.37) | 0.96 |
| APRI | 248 (4.32) | 5498 (95.68) | -0.37 | 0.69 (0.44-1.10) | 0.12 | -0.30 | 0.74 (0.48-1.14) | 0.17 |
| BARD | 1465 (25.43) | 4295 (74.57) | 0.27 | 1.30 (0.90-1.90) | 0.17 | 0.28 | 1.33 (0.92-1.93) | 0.13 |

Abbreviations: FIB-4, Fibrosis-4; NFS, NAFLD fibrosis score; APRI, aspartate aminotransferase to platelets ratio index; SLD, steatotic liver disease; LF, liver fibrosis; aOR, adjusted odds ratio; AHM, antihypertensive medication; IPTW, inverse probability of treatment weighting.

**Supplemental Table 6. Adjusted logistic regression models for the association between antihypertensive medication (AHM) use and liver fibrosis (LF) in individuals with MASLD defined by HSI**

| | | | IPTW and Sampling weighting | | | Only Sampling Weighting | | |
|---|---|---|---|---|---|---|---|---|
| Outcomes | Cases (LF) NO. (%) | Controls (non-LF) No. (%) | Logistic regression coefficient | aOR (95% CI) | P-value | Logistic regression coefficient | aOR (95% CI) | P-value |
| FIB-4 | 11 (0.93) | 1171 (99.07) | -0.51 | 0.60 (0.13-2.81) | 0.52 | -0.47 | 0.62 (0.15-2.54) | 0.15 |
| NFS | 453 (25.42) | 1329 (74.58) | 0.45 | 1.57 (0.88-2.81) | 0.12 | 0.40 | 1.49 (0.89-2.53) | 0.89 |
| APRI | 65 (3.65) | 1717 (96.35) | -0.02 | 0.98 (0.40-2.45) | 0.97 | -0.12 | 0.88 (0.37-2.13) | 0.37 |
| BARD | 434 (24.23) | 1347 (75.77) | -0.14 | 0.87 (0.47-1.60) | 0.65 | 0.04 | 1.04 (0.57-1.89) | 0.57 |

Abbreviations: FIB-4, Fibrosis-4; NFS, NAFLD fibrosis score; APRI, aspartate aminotransferase to platelets ratio index; MASLD, metabolic dysfunction-associated steatotic liver disease; LF, liver fibrosis; aOR, adjusted odds ratio; AHM, antihypertensive medication; IPTW, inverse probability of treatment weighting.

**Supplemental Table 7. Predictive performance of logistic models using FIB-4, NFS, APRI, and BARD for VCTE-defined fibrosis**

| Predictor of Liver Fibrosis | AUROC (%) | PPV at Optimal Threshold (%) | NPV at Optimal Threshold (%) |
|---|---|---|---|
| FIB-4 | 60.91 (58.45-63.36) | 27.80 | 91.39 |
| NFS | 74.06 (71.83-76.29) | 24.58 | 93.72 |
| APRI | 60.75 (58.01-63.49) | 27.82 | 91.72 |
| BARD | 73.97 (72.04-75.89) | 26.31 | 95.28 |

Note: PPV (Positive Predictive Value) = Ture Positive/ (True Positive + False Positive); NPV (Negative Predictive Value) = True Negative/ (True Negative + False Negative). The threshold represents the probability used to determine whether a person is classified as having liver fibrosis based on the predicted probability from the fitted logistic regression model. Each logistic regression model incorporates only one of the predictors above. The optimal threshold is identified as the one corresponding to the highest Youden's J statistic among all evaluated thresholds.

Abbreviations: AUROC, area under receiving operating curves; FIB-4, Fibrosis-4; NFS, NAFLD fibrosis score; APRI, aspartate aminotransferase to platelets ratio index

**Supplemental Table 8. Adjusted Cox models for all-cause mortality in SLD and MASLD defined by US-FLI and FLI**

| Non-invasive markers | | | | | **IPTW and Sampling Weighting** | | **Only Sampling Weighting** | |
|---|---|---|---|---|---|---|---|---|
| | | **Patients NO.(%)** | **Event NO.(%)** | **PYs** | **aHR (95% CI)** | **P-value** | **aHR (95% CI)** | **P-value** |
| US-FLI | No SLD | 14482 (65.57) | 1818 (55.77) | 144640.10 | *Reference* | | *Reference* | |
| | SLD | 7606 (34.43) | 1442 (44.23) | 70286.17 | 1.10 | 0.40 | 0.99 | 0.88 |
| | No MASLD | 16716 (88.12) | 2078 (83.32) | 165947.20 | *Reference* | | *Reference* | |
| | MASLD | 2253 (11.88) | 416 (16.68) | 20573.58 | 0.86 | 0.16 | 0.87 | 0.09 |
| FLI | ≤ 30 | 7253 (42.97) | 843 (35.57) | 72708.67 | *Reference* | | *Reference* | |
| | ≥60 | 9625 (57.03) | 1527 (64.43) | 91372.08 | 0.32 | 0.62 | 0.86 | 0.12 |

Abbreviations: SLD, steatotic liver disease; MASLD, metabolic dysfunction-associated steatotic liver disease; aHR, adjusted hazard ratio; PYs, person-years; IPTW, inverse probability of treatment weighting.

**Supplemental Table 9. Adjusted Cox models for cardiovascular mortality in individuals with SLD**

| AHM-class | (SLD, CVD mortality) | | | | | | |
|---|---|---|---|---|---|---|---|
| | IPTW and sampling weighting | | | | | Only sampling weighting | |
| | Patients NO.(%) | Event NO.(%) | PYs | aHR (95% CI) | P-value | aHR (95% CI) | P-value |
| CCB | 642 (13.65) | 80 (13.94) | 4801.33 | *Reference* | | *Reference* | |
| ACEI | 1273 (27.07) | 134 (23.34) | 10593.42 | 0.67 (0.26,1.73) | 0.40 | 0.52(0.21,1.30) | 0.16 |
| Beta blocker | 1192 (25.35) | 163 (28.40) | 9079.92 | 1.51 (0.64,3.52) | 0.34 | 1.14(0.52,2.49) | 0.75 |
| Diuretic | 1043 (22.18) | 149 (25.96) | 8397.42 | 1.00 (0.41,2.43) | 0.10 | 0.87(0.38,1.99) | 0.74 |
| ARBs | 552 (11.74) | 48 (8.36) | 4077.33 | 1.14 (0.41,3.13) | 0.81 | 0.78(0.30,2.04) | 0.62 |

Abbreviations: CVD, cardiovascular disease; SLD, steatotic liver disease; ACEI, angiotensin-converting enzyme inhibitor; ARB, angiotensin receptor blocker; CCB, calcium channel blocker; aHR, adjusted hazard ratio; AHM, antihypertensive medication; PYs, person-years; IPTW, inverse probability of treatment weighting.

**Supplemental Table 10. Adjusted Cox models for cardiovascular mortality in individuals with MASLD**

| AHM-class | (MASLD, CVD mortality) | | | | | | |
|---|---|---|---|---|---|---|---|
| | IPTW and sampling weighting | | | | | Only sampling weighting | |
| | Patients NO.(%) | Event NO.(%) | PYs | aHR (95% CI) | P-value | aHR (95% CI) | P-value |
| CCB | 208 (13.45) | 22(11.76) | 1442.83 | *Reference* | | *Reference* | |
| ACEI | 414 (26.76) | 43(22.99) | 3430.08 | 0.53 (0.18,1.52) | 0.19 | 0.43 (0.16,1.13) | 0.09 |
| Beta blocker | 413 (26.70) | 61(32.62) | 3192.42 | 1.45 (0.44,4.76) | 0.52 | 1.22 (0.32,4.72) | 0.77 |
| Diuretic | 337 (21.78) | 49(26.20) | 2646.67 | 1.02 (0.20,5.16) | 0.99 | 0.39 (0.05,3.30) | 0.39 |
| ARB | 175 (11.31) | 12(6.42) | 1397.83 | 0.05 (0.00,1.42) | 0.07 | 0.07 (0.00,1.84) | 0.11 |

Abbreviations: CVD, cardiovascular disease; MASLD, metabolic dysfunction-associated steatotic liver disease; ACEI, angiotensin-converting enzyme inhibitor; ARB, angiotensin receptor blocker; CCB, calcium channel blocker; aHR, adjusted hazard ratio; AHM, antihypertensive medication; PYs, person-years; IPTW, inverse probability of treatment weighting.

**Supplemental Table 11. Subgroup analysis by sex: associations between AHM class and all-cause mortality in MASLD**

| (MASLD, All-cause mortality) | | | | | | |
|---|---|---|---|---|---|---|
| **Subgroup** | **AHM-class** | **Patients NO.(%)** | **Event NO.(%)** | **PYs** | **aHR(95%CI)** | |
| Men | CCB | 138 (13.19) | 49 (13.69) | 924.50 | Reference | |
| | ACEI | 293 (28.01) | 88 (24.58) | 2350.00 | 0.27 (0.08-0.90) | |
| | Beta blocker | 290 (27.72) | 109 (30.45) | 2221.92 | 0.53 (0.16-1.78) | |
| | Diuretic | 210 (20.08) | 83 (23.18) | 1593.00 | 0.38 (0.09-1.56) | |
| | ARB | 115 (10.99) | 29 (8.10) | 881.00 | 0.36 (0.07-1.87) | |
| | | | | | **aHR (95%CI)** | **P-value interaction** |
| Women | CCB | 70 (11.69) | 20 (26.32) | 518.33 | Reference | |
| | ACEI | 121 (26.07) | 27 (31.58) | 1080.08 | 0.60 (0.13-2.76) | 0.42 |
| | Beta blocker | 123 (24.94) | 29 (27.91) | 970.50 | 0.25 (0.04-1.80) | 0.54 |
| | Diuretic | 127 (26.29) | 35 (36.84) | 1053.67 | 0.74 (0.13-4.22) | 0.56 |
| | ARB | 60 (11.01) | 8 (5.26) | 516.83 | 0.10 (0.01-1.26) | 0.41 |

Note: The result is obtained from Adjusted Cox proportional hazards models with IPTW and sampling weighting.

The HR for women taking a specific medication class is determined by combining the coefficient for the main effect of the medication class with the coefficient for the interaction between the medication class and female gender.

Abbreviations: MASLD, metabolic dysfunction-associated steatotic liver disease; ACEI, angiotensin-converting enzyme inhibitor; ARB, angiotensin receptor blocker; CCB, calcium channel blocker; aHR, adjusted hazard ratio; AHM, antihypertensive medication; PYs, person-years; IPTW, inverse probability of treatment weighting.

**Supplemental Table 12. Subgroup analysis by income: associations between AHM class and all-cause mortality in SLD**

| (SLD, All-cause mortality) | | | | | | |
|---|---|---|---|---|---|---|
| **Subgroup** | **AHM-class** | **Patients NO.(%)** | **Event NO.(%)** | **PYs** | **aHR (95%CI)** | **P-value** |
| | CCB | 113 (33.95) | 28 (33.62) | 827.00 | *Reference* | |
| PIR<= 1 | ACEI | 237 (28.08) | 78 (28.02) | 1911.33 | 1.16 (0.40-3.39) | 0.78 |
| | Beta blocker | 196 (24.17) | 65 (25.00) | 1423.67 | 0.94 (0.20-4.51) | 0.94 |
| | Diuretic | 182 (26.07) | 66 (28.45) | 1394.50 | 0.72 (0.20-2.63) | 0.62 |
| | ARB | 83 (11.89) | 23 (9.91) | 566.42 | 2.65 (0.75-9.29) | 0.13 |
| | | | | | **aHR (95%CI)** | **P-value interaction** |
| | CCB | 342 (31.36) | 128 (27.96) | 2606.25 | *Reference* | |
| 1< PIR< 4 | ACEI | 663 (29.38) | 215 (30.43) | 5268.33 | 0.50 (0.24-1.06) | 0.20 |
| | Beta blocker | 621 (25.29) | 234 (26.09) | 4538.58 | 0.80 (0.37-1.74) | 0.86 |
| | Diuretic | 568 (26.87) | 235 (30.56) | 4511.75 | 1.17 (0.50-2.73) | 0.53 |
| | ARB | 262 (12.39) | 85 (11.05) | 1917.75 | 0.72 (0.32-1.63) | 0.08 |
| | | | | | **aHR(95%CI)** | **P-value interaction** |
| | CCB | 119 (28.52) | 31 (14.83) | 943.58 | *Reference* | |
| PIR>= 4 | ACEI | 253 (29.76) | 45 (21.53) | 2554.08 | 0.35 (0.09-1.36) | 0.18 |
| | Beta blocker | 264 (26.24) | 60 (28.71) | 2288.00 | 0.30 (0.07-1.33) | 0.29 |
| | Diuretic | 205 (23.11) | 45 (21.53) | 1799.25 | 0.47 (0.12-1.86) | 0.68 |
| | ARB | 165 (18.60) | 28 (13.40) | 1317.92 | 0.50 (0.11-2.23) | 0.10 |

Note: The result is obtained from adjusted Cox proportional hazards models with IPTW and sampling weighting.

Abbreviations: SLD, steatotic liver disease; PIR, family income-to-poverty ratio; ACEI, angiotensin-converting enzyme inhibitor; ARB, angiotensin receptor blocker; CCB, calcium channel blocker; aHR, adjusted hazard ratio; AHM, antihypertensive medication; PYs, person-years; IPTW, inverse probability of treatment weighting.

**Supplemental Table 13. Subgroup analysis by diabetes status: associations between AHM class and all-cause mortality in MASLD**

| (MASLD, All-cause mortality) | | | | | | |
|---|---|---|---|---|---|---|
| **Subgroup** | **AHM-class** | **Patients NO.(%)** | **Event NO.(%)** | **PYs** | **aHR (95%CI)** | **P-value** |
| No diabetes | CCB | 96 (12.34) | 35 (15.22) | 760.58 | *Reference* | |
| | ACEI | 192 (24.68) | 53 (23.04) | 1706.67 | 0.35 (0.11-1.10) | 0.07 |
| | Beta blocker | 218 (28.02) | 67 (29.13) | 1853.83 | 0.39 (0.13-1.21) | 0.10 |
| | Diuretic | 177 (22.75) | 57 (24.78) | 1549.92 | 0.42 (0.13-1.36) | 0.15 |
| | ARB | 95 (12.21) | 18 (7.83) | 841.50 | 0.15 (0.02-1.07) | 0.06 |
| | | | | | **aHR (95%CI)** | **P-value interaction** |
| Diabetes | CCB | 112 (14.56) | 34 (13.77) | 682.25 | *Reference* | |
| | ACEI | 222 (28.87) | 62 (25.10) | 1723.42 | 0.31 (0.07-1.36) | 0.89 |
| | Beta blocker | 195 (25.36) | 71 (25.36) | 1338.58 | 0.69 (0.14-3.26) | 0.55 |
| | Diuretic | 160 (20.81) | 61 (20.81) | 1096.75 | 0.63 (0.07-5.45) | 0.73 |
| | ARB | 80 (10.40) | 19 (10.40) | 556.33 | 0.78 (0.19-3.26) | 0.16 |

Note: The result is obtained from adjusted Cox proportional hazards models with IPTW and sampling weighting.

Abbreviations: MASLD, metabolic dysfunction-associated steatotic liver disease; ACEI, angiotensin-converting enzyme inhibitor; ARB, angiotensin receptor blocker; CCB, calcium channel blocker; aHR, adjusted hazard ratio; AHM, antihypertensive medication; PYs, person-years. IPTW, inverse probability of treatment weighting.

**Supplemental Table 14. Sensitivity analysis in SLD: using alternative reference groups to compare AHM classes and all-cause mortality**

| Baseline | ACEI | | Beta blocker | | Diuretic | | ARB | |
|---|---|---|---|---|---|---|---|---|
| **AHM-class** | **aHR (95% CI)** | **P-value** | **aHR (95% CI)** | **P-value** | **aHR (95% CI)** | **P-value** | **aHR (95% CI)** | **P-value** |
| ACEI | *Reference* | | 0.82 (0.50-1.33) | 0.42 | 0.61 (0.35-1.06) | 0.08 | 0.67 (0.40-1.13) | 0.13 |
| Beta blocker | 1.22 (0.75-1.99) | 0.42 | *Reference* | | 0.75 (0.41-1.35) | 0.34 | 0.82 (0.48-1.40) | 0.48 |
| Diuretic | 1.64 (0.94-2.84) | 0.08 | 1.34 (0.74-2.43) | 0.34 | *Reference* | | 1.10 (0.61-1.99) | 0.75 |
| ARB | 1.48 (0.89-2.48) | 0.13 | 1.21 (0.71-2.07) | 0.48 | 0.91 (0.50-1.64) | 0.75 | *Reference* | |
| CCB | 1.73 (0.97-3.07) | 0.06 | 1.42 (0.77-2.62) | 0.27 | 1.06 (0.56-2.00) | 0.86 | 1.17 (0.63-2.16) | 0.62 |

Note: The result is obtained from adjusted Cox proportional hazards models with IPTW and sampling weighting.

Abbreviations: SLD, steatotic liver disease; ACEI, angiotensin-converting enzyme inhibitor; ARB, angiotensin receptor blocker; CCB, calcium channel blocker; aHR, adjusted hazard ratio; AHM, antihypertensive medication; IPTW, inverse probability of treatment weighting.

**Supplemental Table 15. Sensitivity analysis in MASLD: using alternative IPTW models to evaluate associations between AHM class and all-cause mortality**

| | (MASLD, All-cause mortality) | |
|---|---|---|
| **AHM-class** | **aHR (95% CI)** | **P-value** |
| CCB | *Reference* | |
| ACEI | 0.31 (0.12-0.85) | 0.02 |
| Beta blocker | 0.38 (0.14-1.06) | 0.07 |
| Diuretic | 0.47 (0.16-1.43) | 0.18 |
| ARB | 0.25 (0.06-0.98) | 0.046 |

Note: The result is obtained from adjusted Cox proportional hazards models with IPTW and sampling weighting.

Abbreviations: MASLD, metabolic dysfunction-associated steatotic liver disease; ACEI, angiotensin-converting enzyme inhibitor; ARB, angiotensin receptor blocker; CCB, calcium channel blocker; aHR, adjusted hazard ratio; AHM, antihypertensive medication; IPTW, inverse probability of treatment weighting.
